# Long-Term Reintervention, Clinical Valve Failure, and Outcomes After Transcatheter Aortic Valve Replacement: A National Real-World Study

**DOI:** 10.64898/2026.08.16.26360543

**Authors:** Zhiyuan Ma, Cyrus P Elmi, Steven M Stevens, Amit Gupta, Peter Puleo, Jamshid Shirani

## Abstract

**Background:** As transcatheter aortic valve replacement (TAVR) expands to younger patients with longer life expectancy, understanding long-term reintervention and clinically significant valve failure has become increasingly important.

**Objectives:** To evaluate temporal trends in TAVR outcomes, characterize the incidence and timing of aortic valve reintervention, compare outcomes after redo-TAVR (TAVR-in-TAVR) versus surgical explantation, and assess freedom from clinically significant valve failure requiring repeat intervention after TAVR versus surgical bioprosthetic aortic valve replacement (SAVR).

**Methods:** We performed a retrospective cohort study using the Epic Cosmos. Adults undergoing index TAVR between February 2010 and May 2026 were identified. Primary outcomes included aortic valve reintervention and 30-day major adverse cardiovascular events (MACE). Reintervention incidence was estimated using competing-risk methods with death as the competing event. Propensity-score matching compared redo-TAVR with surgical explantation and TAVR with SAVR. A prespecified 1-year landmark analysis evaluated clinically significant valve failure requiring repeat intervention.

**Results:** Among 300,927 patients undergoing TAVR, annual procedural volume increased more than tenfold between 2016 and 2025. Thirty-day MACE decreased from 31.8% before 2017 to 18.6% after 2022 (P<0.001), while mortality declined from 3.0% to 1.4% (P<0.001). During follow-up, 3,315 patients underwent redo-TAVR and 347 underwent surgical explantation. The cumulative incidence of reintervention was 1.1%, 1.2%, 1.5%, and 2.7% at 3, 5, 7, and 10 years, respectively, with significantly lower rates in contemporary procedural eras (Gray test, P<0.001). Compared with surgical explantation, redo-TAVR was associated with lower 30-day mortality, stroke, acute kidney injury, major bleeding, and shorter hospitalization. However, among propensity-matched hospital survivors, surgical explantation was associated with superior long-term survival (hazard ratio: 0.64; 95% CI: 0.44–0.93; P=0.018). In the landmark analysis, clinically significant valve failure requiring repeat intervention occurred earlier after TAVR than after SAVR despite a lower overall cumulative incidence of repeat intervention following TAVR.

**Conclusions:** Contemporary TAVR is associated with progressively improving procedural outcomes and a low incidence of repeat aortic valve intervention. Redo-TAVR offers lower perioperative risk than surgical explantation, whereas surgical explantation is associated with superior long-term survival among selected patients. Earlier clinically significant valve failure requiring repeat intervention after TAVR underscores the importance of lifetime management strategies as TAVR expands to younger populations.

## Introduction

Transcatheter aortic valve replacement (TAVR) has transformed the treatment of severe aortic stenosis and is now an established therapy across the full spectrum of surgical risk (1–4). Advances in transcatheter valve technology, procedural techniques, and operator experience have resulted in marked improvements in procedural safety and short-term clinical outcomes (5), supporting the expansion of TAVR into younger patients with longer anticipated survival. As the population of long-term TAVR survivors grows, attention has shifted from procedural success to lifetime management. Repeat aortic valve intervention represents one of the most consequential late complications because it often reflects clinically significant bioprosthetic valve failure and requires complex decision-making regarding redo transcatheter intervention or surgical explantation. Although redo-TAVR offers a less invasive treatment strategy, its feasibility may be limited by unfavorable anatomy, coronary access, valve configuration, or the mechanism of prosthetic valve failure. Surgical explantation provides definitive treatment but is technically challenging and associated with greater procedural risk. Despite increasing clinical interest, contemporary real-world data regarding the incidence, timing, durability and outcomes of aortic valve reintervention after TAVR remain limited (6–10). Furthermore, comparative data evaluating clinically significant valve failure requiring repeat intervention after transcatheter versus surgical bioprosthetic valve replacement are sparse.

Using the Epic Cosmos national electronic health record network, we sought to characterize temporal trends in TAVR utilization and early clinical outcomes, determine the incidence and timing of aortic valve reintervention after TAVR, compare clinical outcomes following redo-TAVR and surgical explantation, and evaluate freedom from clinically significant valve failure requiring repeat intervention following transcatheter and surgical bioprosthetic aortic valve replacement.

## METHODS

### Data Source

We conducted a retrospective cohort study using the Epic Cosmos Expertly Determined De-Identified (EDDI) database, a dataset created in collaboration with a community of health systems representing more than 304 million patients receiving care across participating U.S. health systems. Adult patients undergoing TAVR or surgical aortic valve replacement (SAVR) between February 2010 and May 2026 were identified. The study adhered to the Declaration of Helsinki and the Strengthening the Reporting of Observational Studies in Epidemiology (STROBE) reporting guideline. Because all data were fully de-identified, the study was exempt from institutional review board (IRB) review (IRB-FY2026-117).

### Study Population

Adults aged ≥18 years undergoing index TAVR or SAVR during the study period were eligible for inclusion. Patients with implausible body mass index values (<15 or >100 kg/m²) or an invalid death date preceding the index procedure were excluded. Aortic valve reintervention was defined as any repeat transcatheter or surgical aortic valve procedure performed at least one calendar day after the index TAVR or SAVR. Because Epic Cosmos records procedures by calendar date rather than time of day, same day repeat procedures could not be distinguished from the index intervention and were therefore excluded.

### Study Definitions

Baseline comorbidities were identified using diagnosis codes recorded before the index hospitalization. The primary composite endpoint, major adverse cardiovascular events (MACE), consisted of all-cause mortality, stroke, permanent pacemaker implantation, major bleeding, and acute kidney injury occurring within 30 days after TAVR. Procedures including TAVR, SAVR, and permanent pacemaker implantation were identified using ICD-10-PCS and Current Procedural Terminology (CPT) codes (Supplemental Table 1). Comorbidities and postoperative complications were identified using ICD-10 diagnosis codes. Patients were followed from the index procedure until death or May 13, 2026, whichever occurred first.

### Outcomes

The primary outcomes were incidence of aortic valve reintervention after index TAVR and 30-day MACE. Secondary outcomes included timing of reintervention, short-and long-term outcomes following redo-TAVR (TAVR-in-TAVR) versus surgical explantation, and comparative freedom from clinically significant valve failure requiring repeat intervention following TAVR and SAVR.

### Assessment of Clinically Significant Valve Failure

Because Epic Cosmos does not contain longitudinal echocardiographic or computed tomography data necessary to directly identify structural valve deterioration or bioprosthetic valve dysfunction, clinically significant valve failure was assessed using repeat aortic valve intervention as a pragmatic clinical surrogate endpoint. For TAVR recipients, time to clinically significant valve failure was defined as the interval from the index TAVR procedure to redo-TAVR or surgical explantation. For SAVR recipients, time was defined as the interval from the index SAVR procedure to repeat SAVR or valve-in-valve TAVR.

Because reinterventions during the first postoperative year are more likely to reflect procedural complications than intrinsic valve degeneration, we prespecified a 1-year landmark analysis. Repeat interventions occurring beyond one year were considered to represent probable clinically significant bioprosthetic valve failure requiring reintervention and constituted the primary cohort for comparative durability analyses. Freedom from clinically significant valve failure requiring repeat intervention was estimated using Kaplan-Meier methods and quantified with restricted mean survival time (RMST), representing the average duration free from repeat intervention over follow-up intervals.

### Statistical Analysis

Continuous variables are presented as mean (95% confidence interval) or median (interquartile range), as appropriate, and categorical variables as frequency (percentage). Comparisons were performed using Student’s t test or the Mann-Whitney U test for continuous variables and the chi-square test for categorical variables.

Because death precludes observation of valve reintervention, cumulative incidence was estimated using competing-risk methods treating all-cause mortality as the competing event. Cumulative incidence functions were compared using Gray’s test (11). Overall survival was estimated using the Kaplan-Meier method and compared with the log-rank test. Cox proportional hazards models were used to estimate hazard ratios (HRs) with 95% confidence intervals (CIs). To identify predictors of clinically significant valve failure requiring repeat intervention beyond one year, baseline characteristics were first evaluated by univariable analysis. Variables demonstrating statistical significance together with clinically relevant covariates were entered into multivariable logistic regression models to estimate adjusted odds ratios (ORs) and 95% CIs.

To reduce treatment-selection bias, propensity-score matching was performed separately for redo-TAVR versus surgical explantation and TAVR versus SAVR. Propensity scores were estimated using multivariable logistic regression incorporating demographic characteristics, cardiovascular risk factors, major comorbidities, previous cardiovascular procedures, implantable cardiac devices, year of intervention, body mass index, and Charlson Comorbidity Index, as appropriate for each comparison. Patients were matched 1:1 using nearest-neighbor matching with a caliper width of 0.05. RMST was calculated to quantify absolute differences in freedom from repeat intervention over prespecified follow-up intervals. All analyses were performed using R version 4.1.2 (R Foundation for Statistical Computing, Vienna, Austria). All tests were two-sided, and P<0.05 was considered statistically significant.

## RESULTS

### Study Population

Between February 2010 and May 2026, 300,927 patients underwent index TAVR and 45,539 underwent SAVR with bioprosthesis within the Epic Cosmos database (Figure 1). Annual TAVR volume increased more than tenfold, from 4,897 procedures in 2016 to 53,668 procedures in 2025 (Supplemental Figure 1). Compared with SAVR recipients, patients undergoing TAVR were substantially older (mean age, 78.9 vs. 67.2 years; P<0.001) and had a greater burden of cardiovascular comorbidity, including diabetes mellitus, hypertension, coronary artery disease, heart failure, chronic kidney disease, prior myocardial infarction, and prior stroke or transient ischemic attack (all P<0.001). In contrast, bicuspid aortic valve disease was more common among patients undergoing SAVR (28.1% vs. 6.3%; P<0.001). Baseline characteristics are summarized in Table 1.

**Figure 1.**
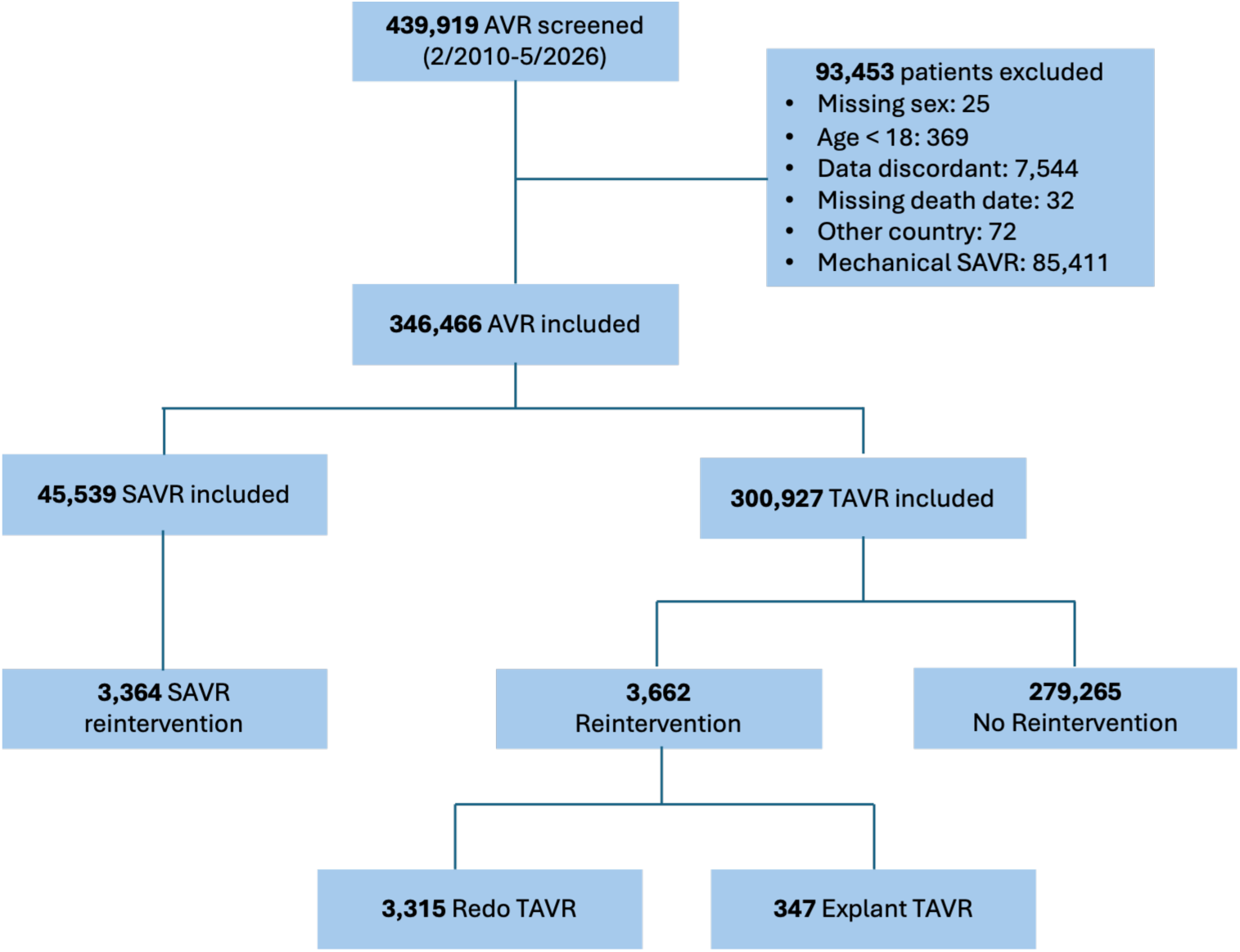
Study flow chart. SAVR = surgical aortic valve replacement; TAVR = transcatheter aortic valve replacement.

**Table 1.**
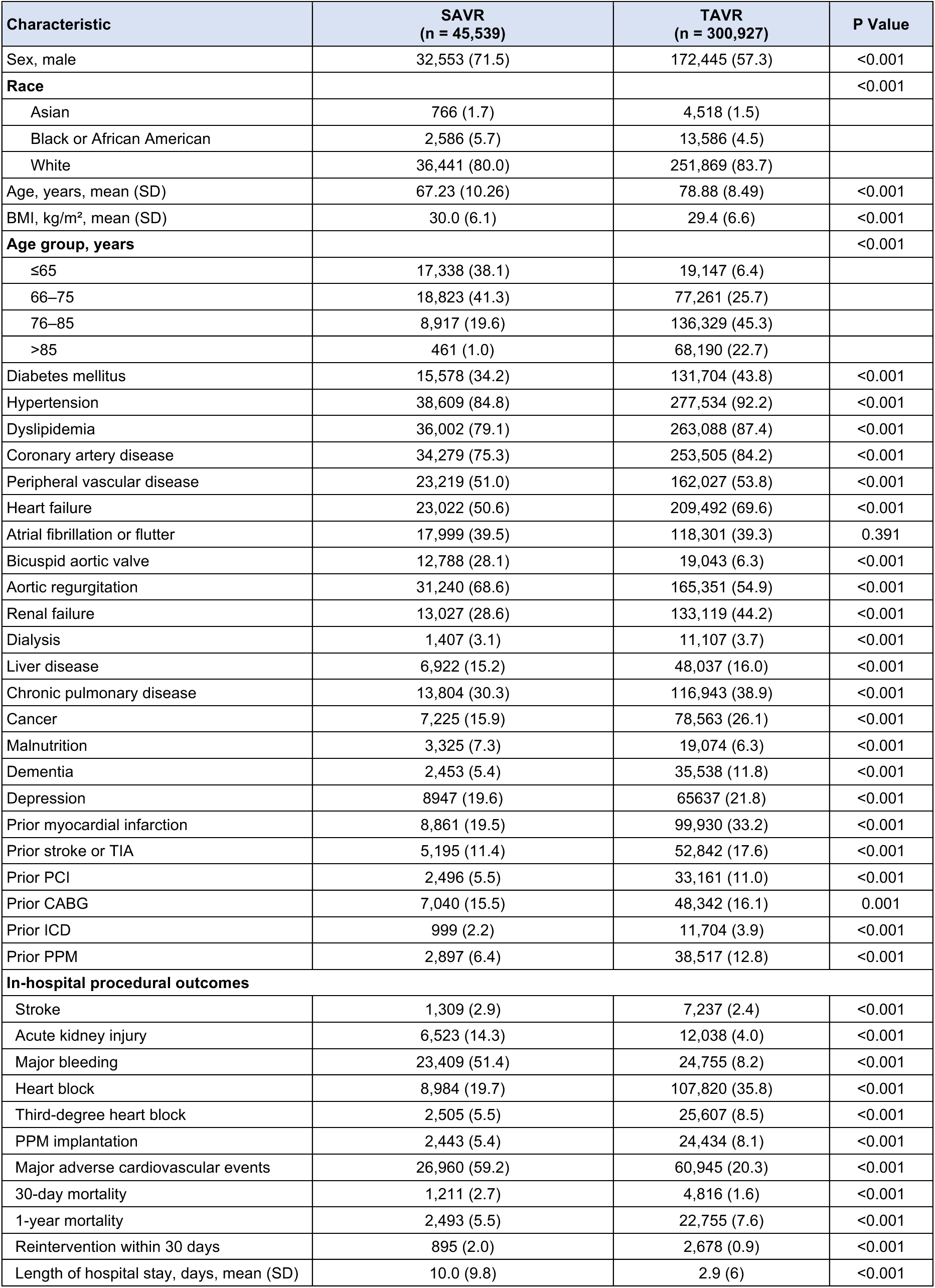
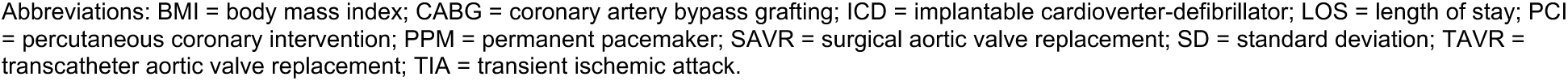
Baseline Characteristics and Clinical Outcomes of Patients Undergoing SAVR) or TAVR.

| Characteristic | SAVR<br>(n = 45,539) | TAVR<br>(n = 300,927) | P Value |
| --- | --- | --- | --- |
| Sex, male | 32,553 (71.5) | 172,445 (57.3) | <0.001 |
| **Race** |  |  | <0.001 |
| Asian | 766 (1.7) | 4,518 (1.5) |  |
| Black or African American | 2,586 (5.7) | 13,586 (4.5) |  |
| White | 36,441 (80.0) | 251,869 (83.7) |  |
| Age, years, mean (SD) | 67.23 (10.26) | 78.88 (8.49) | <0.001 |
| BMI, kg/m<sup>2</sup>, mean (SD) | 30.0 (6.1) | 29.4 (6.6) | <0.001 |
| **Age group, years** |  |  | <0.001 |
| ≤65 | 17,338 (38.1) | 19,147 (6.4) |  |
| 66–75 | 18,823 (41.3) | 77,261 (25.7) |  |
| 76–85 | 8,917 (19.6) | 136,329 (45.3) |  |
| >85 | 461 (1.0) | 68,190 (22.7) |  |
| Diabetes mellitus | 15,578 (34.2) | 131,704 (43.8) | <0.001 |
| Hypertension | 38,609 (84.8) | 277,534 (92.2) | <0.001 |
| Dyslipidemia | 36,002 (79.1) | 263,088 (87.4) | <0.001 |
| Coronary artery disease | 34,279 (75.3) | 253,505 (84.2) | <0.001 |
| Peripheral vascular disease | 23,219 (51.0) | 162,027 (53.8) | <0.001 |
| Heart failure | 23,022 (50.6) | 209,492 (69.6) | <0.001 |
| Atrial fibrillation or flutter | 17,999 (39.5) | 118,301 (39.3) | 0.391 |
| Bicuspid aortic valve | 12,788 (28.1) | 19,043 (6.3) | <0.001 |
| Aortic regurgitation | 31,240 (68.6) | 165,351 (54.9) | <0.001 |
| Renal failure | 13,027 (28.6) | 133,119 (44.2) | <0.001 |
| Dialysis | 1,407 (3.1) | 11,107 (3.7) | <0.001 |
| Liver disease | 6,922 (15.2) | 48,037 (16.0) | <0.001 |
| Chronic pulmonary disease | 13,804 (30.3) | 116,943 (38.9) | <0.001 |
| Cancer | 7,225 (15.9) | 78,563 (26.1) | <0.001 |
| Malnutrition | 3,325 (7.3) | 19,074 (6.3) | <0.001 |
| Dementia | 2,453 (5.4) | 35,538 (11.8) | <0.001 |
| Depression | 8947 (19.6) | 65637 (21.8) | <0.001 |
| Prior myocardial infarction | 8,861 (19.5) | 99,930 (33.2) | <0.001 |
| Prior stroke or TIA | 5,195 (11.4) | 52,842 (17.6) | <0.001 |
| Prior PCI | 2,496 (5.5) | 33,161 (11.0) | <0.001 |
| Prior CABG | 7,040 (15.5) | 48,342 (16.1) | 0.001 |
| Prior ICD | 999 (2.2) | 11,704 (3.9) | <0.001 |
| Prior PPM | 2,897 (6.4) | 38,517 (12.8) | <0.001 |
| **In-hospital procedural outcomes** |  |  |  |
| Stroke | 1,309 (2.9) | 7,237 (2.4) | <0.001 |
| Acute kidney injury | 6,523 (14.3) | 12,038 (4.0) | <0.001 |
| Major bleeding | 23,409 (51.4) | 24,755 (8.2) | <0.001 |
| Heart block | 8,984 (19.7) | 107,820 (35.8) | <0.001 |
| Third-degree heart block | 2,505 (5.5) | 25,607 (8.5) | <0.001 |
| PPM implantation | 2,443 (5.4) | 24,434 (8.1) | <0.001 |
| Major adverse cardiovascular events | 26,960 (59.2) | 60,945 (20.3) | <0.001 |
| 30-day mortality | 1,211 (2.7) | 4,816 (1.6) | <0.001 |
| 1-year mortality | 2,493 (5.5) | 22,755 (7.6) | <0.001 |
| Reintervention within 30 days | 895 (2.0) | 2,678 (0.9) | <0.001 |
| Length of hospital stay, days, mean (SD) | 10.0 (9.8) | 2.9 (6) | <0.001 |

Abbreviations: BMI = body mass index; CABG = coronary artery bypass grafting; ICD = implantable cardioverter-defibrillator; LOS = length of stay; PCI = percutaneous coronary intervention; PPM = permanent pacemaker; SAVR = surgical aortic valve replacement; SD = standard deviation; TAVR = transcatheter aortic valve replacement; TIA = transient ischemic attack.

### Temporal Trends in Early Clinical Outcomes

Short-term outcomes improved substantially throughout the study period. Patients were stratified into four procedural eras: before 2017, 2017–2019, 2020–2022, and after 2022. The incidence of 30-day MACE declined progressively from 31.8% before 2017 to 25.7%, 19.6%, and 18.6% across successive eras (P<0.001) (Central Illustration and Table 2).

**Table 2.** Thirty-Day Clinical Outcomes Following TAVR According to Procedural Era.

| Thirty-day procedural outcomes | <2017<br>(n = 5,748) | 2017–2019<br>(n = 46,247) | 2020–2022<br>(n = 87,151) | >2022<br>(n = 161,781) | P Value |
| --- | --- | --- | --- | --- | --- |
| Stroke | 151 (2.6) | 1,302 (2.8) | 2,132 (2.4) | 3,652 (2.3) | <0.001 |
| Acute kidney injury | 343 (6.0) | 2,373 (5.1) | 3,334 (3.8) | 5,988 (3.7) | <0.001 |
| Major bleeding | 1,027 (17.9) | 5,732 (12.4) | 6,952 (8.0) | 11,044 (6.8) | <0.001 |
| Heart block | 1,394 (24.3) | 15,041 (32.5) | 29,868 (34.3) | 61,517 (38.0) | <0.001 |
| Third-degree heart block | 505 (8.8) | 3,969 (8.6) | 7,115 (8.2) | 14,018 (8.7) | <0.001 |
| PPM implantation | 626 (10.9) | 4,303 (9.3) | 6,677 (7.7) | 12,828 (7.9) | <0.001 |
| Length of hospital stay, days,<br>mean (SD) | 4.7 (6.1) | 3.6 (10.1) | 2.9 (5.2) | 2.7 (4.6) | <0.001 |
| Major adverse cardiovascular<br>events | 1,830 (31.8) | 11,869 (25.7) | 17,110 (19.6) | 30,136 (18.6) | <0.001 |
| 30-day mortality | 171 (3.0) | 901 (1.9) | 1,475 (1.7) | 2,269 (1.4) | <0.001 |

Abbreviations: SD = standard deviation; TAVR = transcatheter aortic valve replacement.

Marked improvements were observed across nearly all individual complications. Thirty-day mortality decreased by more than one-half (3.0% vs. 1.4%), acute kidney injury declined from 6.0% to 3.7%, major bleeding from 17.9% to 6.8%, and permanent pacemaker implantation from 10.9% to 7.9% (all P<0.001). Stroke rates remained low throughout the study period and decreased modestly in the contemporary era (2.6% vs. 2.3%; P<0.001). Temporal trends are shown in Figure 2.

**Figure 2.**
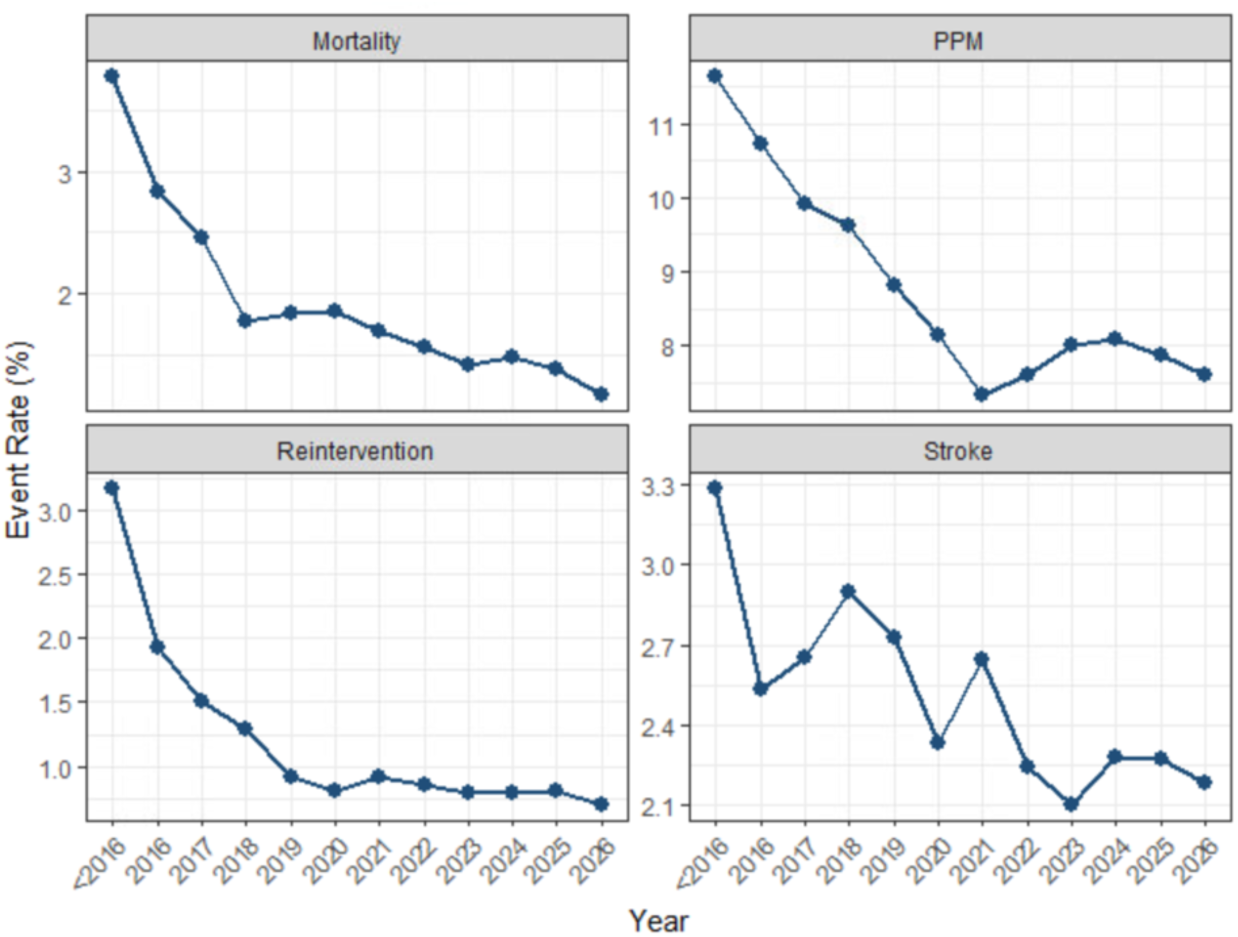
Temporal trends in 30-day clinical outcomes following index transcatheter aortic valve replacement (TAVR), TAVR = transcatheter aortic valve replacement, PPM = permanent pacemaker.

### Incidence and Timing of Aortic Valve Reintervention

Accounting for the competing risk of death, the cumulative incidence of overall aortic valve reintervention remained low, reaching 1.1%, 1.2%, 1.5%, and 2.7% at 3, 5, 7, and 10 years, respectively (Central Illustration and Figure 3A). The incidence of reintervention decreased significantly across successive procedural eras (Gray’s test, P<0.001). Three-year cumulative incidence declined from 3.4% among procedures performed before 2017 to 1.4%, 1.0%, and 0.92% in subsequent eras (Figure 3B).

**Figure 3.**
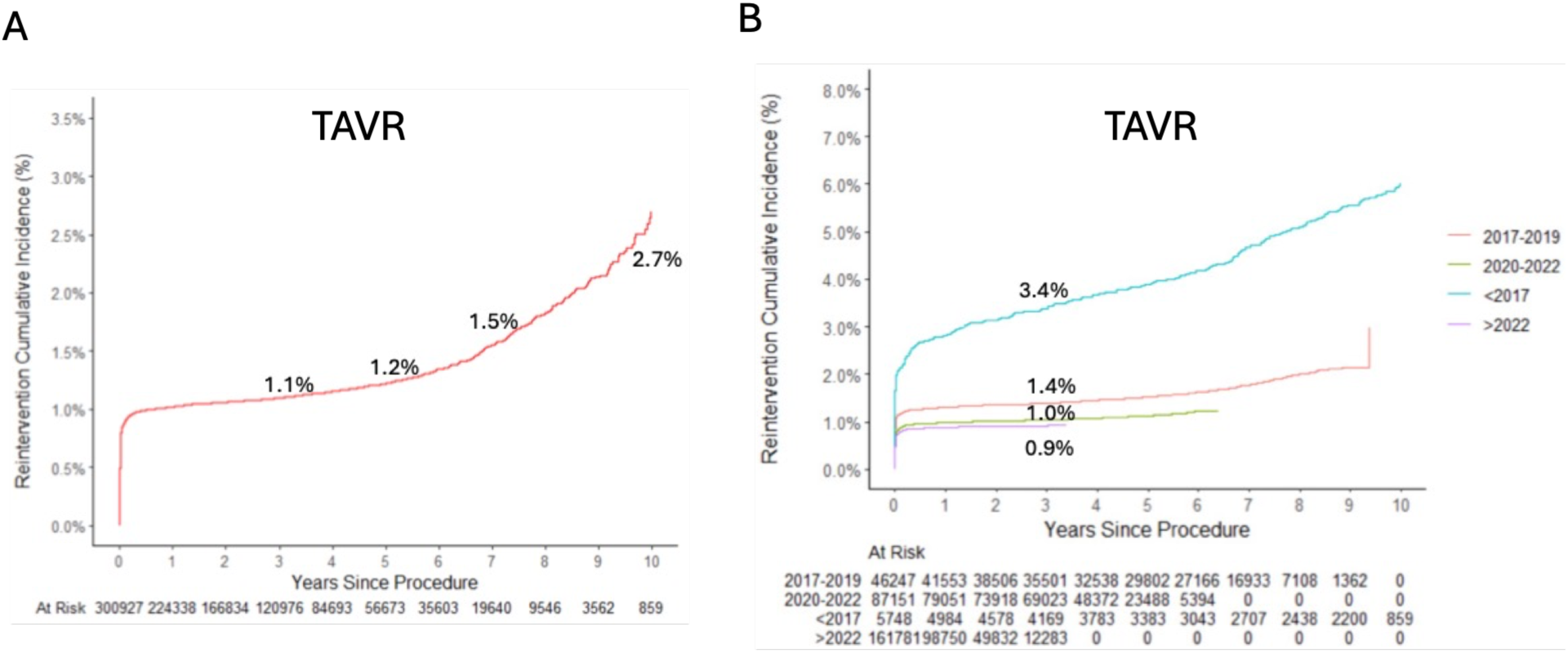
Cumulative incidence of aortic valve reintervention following index transcatheter aortic valve replacement (TAVR). (A) Overall cumulative incidence of aortic valve reintervention after index TAVR. (B) Three-year cumulative incidence of aortic valve reintervention according to the index TAVR procedural era.

In the propensity-score matched cohort comparing patients undergoing index TAVR and surgical bioprosthetic AVR, the cumulative incidence of repeat aortic valve intervention was significantly lower following TAVR than SAVR, reaching 1.3%, 1.6%, 2.4%, and 5.1% at 3, 5, 7, and 10 years, respectively, compared with 3.2%, 4.8%, 8.9%, and 31.0% following SAVR (Gray’s test, P<0.001; Supplemental Figure 2A). With a median follow-up of 3.5 years, Kaplan– Meier analysis demonstrated significantly higher long-term all-cause mortality after TAVR than after SAVR (hazard ratio [HR], 1.78; 95% CI, 1.71–1.85; P<0.001; Supplemental Figure 2B), despite the lower cumulative incidence of repeat intervention after TAVR.

During follow-up, 3,315 patients (1.10%) underwent redo-TAVR and 347 (0.12%) underwent surgical explantation after index TAVR (Supplemental Table 2). The timing of reintervention differed substantially according to treatment strategy. Redo-TAVR occurred much earlier than surgical explantation, with a median interval of 2 days (IQR 1–15) versus 490 days (IQR 62– 1,610), respectively (P<0.001). Consequently, 78.7% of redo-TAVR procedures occurred within 30 days of the index procedure, whereas only 19.9% of surgical explantations occurred during this period. The indications for reintervention also differed significantly between treatment strategies. Endocarditis (4.3% vs. 34.3%) and clinically significant bioprosthetic valve failure (12.1% vs. 49.3%) were substantially less common among patients undergoing redo-TAVR than among those undergoing surgical explantation (both P<0.001). Among patients ultimately requiring repeat intervention, redo-TAVR remained concentrated in the early postprocedural period. By 1, 3, and 5 years after the index procedure, 87%, 90%, and 93% of redo-TAVR procedures had already occurred compared with 44%, 62%, and 79% of surgical explantations (all P<0.001) (Figure 4A).

**Figure 4.**
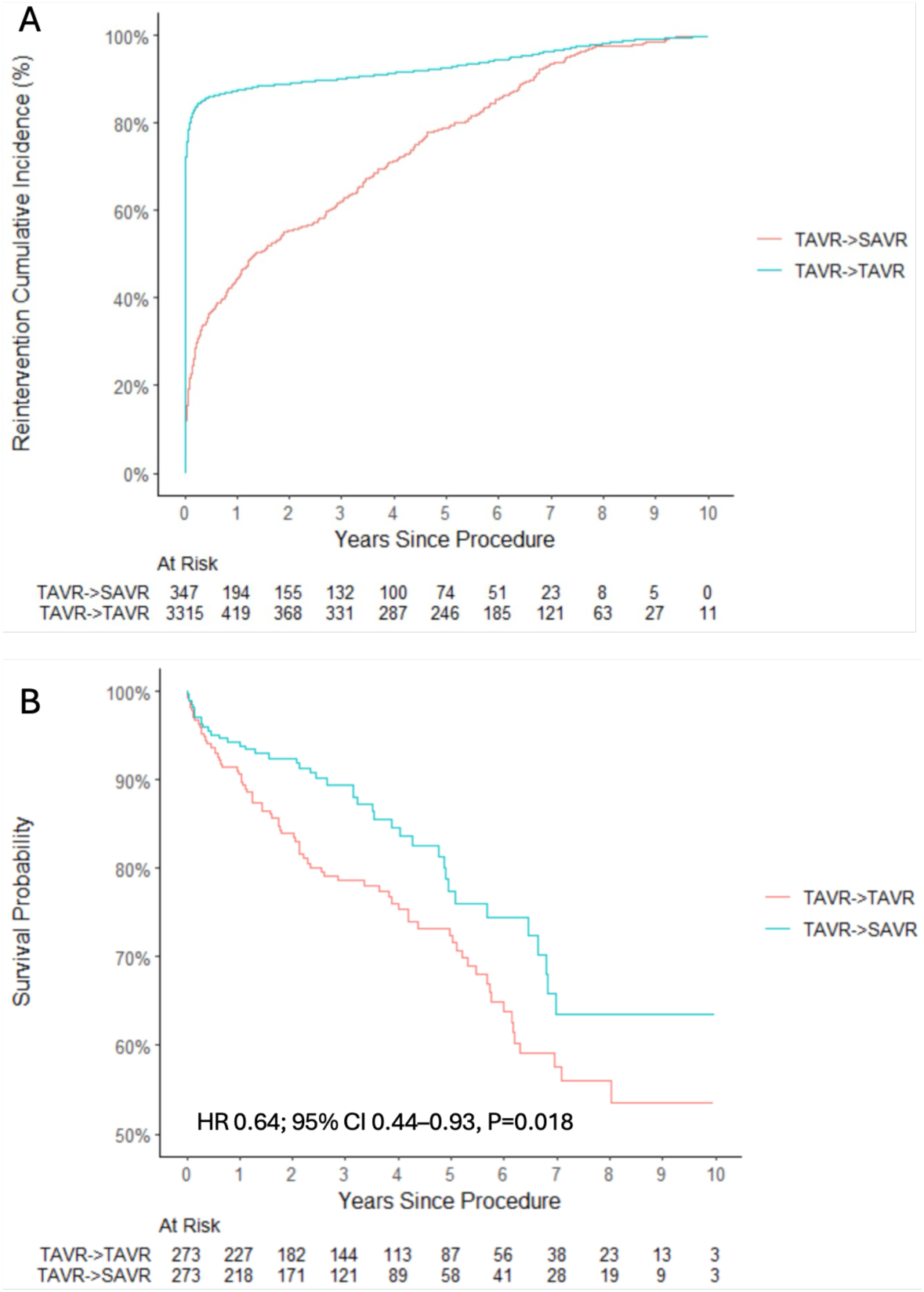
Timing and outcomes of reintervention following index transcatheter aortic valve replacement (TAVR). (A) Cumulative incidence of redo-TAVR and surgical explantation after index TAVR. (B) Kaplan–Meier survival curves among propensity-score–matched hospital survivors undergoing redo-TAVR or surgical explantation.

### Clinical Outcomes Following Reintervention

Patients undergoing redo-TAVR were older than those undergoing surgical explantation both at the index procedure (78.1 vs. 68.4 years; P<0.001) and at reintervention (78.8 vs. 70.9 years; P<0.001) (Supplemental Table 2). Despite their older age, redo-TAVR was associated with substantially better early outcomes than surgical explantation. Thirty-day MACE occurred in 16.8% of patients undergoing redo-TAVR compared with 59.7% after surgical explantation (P<0.001). Compared with surgical explantation, redo-TAVR was associated with significantly lower 30-day mortality (3.2% vs. 8.9%), stroke (1.3% vs. 6.1%), acute kidney injury (3.5% vs. 20.7%), major bleeding (4.8% vs. 41.2%), and permanent pacemaker implantation (7.2% vs. 11.5%) (all P≤0.006). Median length of stay was also substantially shorter following redo-TAVR (3 [IQR 1–8] vs. 12 [IQR 7–20] days; P<0.001).

After propensity-score matching, 273 hospital survivors remained in each treatment group (Supplemental Table 3). With a median follow-up of 3.6 years, surgical explantation was associated with superior long-term survival (HR 0.64; 95% CI 0.44–0.93; P=0.018) (Figure 4B), despite worse perioperative outcomes

### Comparative Clinical Valve Failure Requiring Repeat Intervention

To evaluate clinically significant valve failure, we performed a prespecified 1-year landmark analysis. Of 7,026 patients undergoing repeat intervention, 613 had prior TAVR and 2,208 had prior SAVR more than one year after the index procedure and were included in the landmark cohort (Table 3). Among these patients, the median interval to repeat intervention was significantly shorter following TAVR than SAVR (1,883 vs. 2,279 days; P<0.001).

**Table 3.**
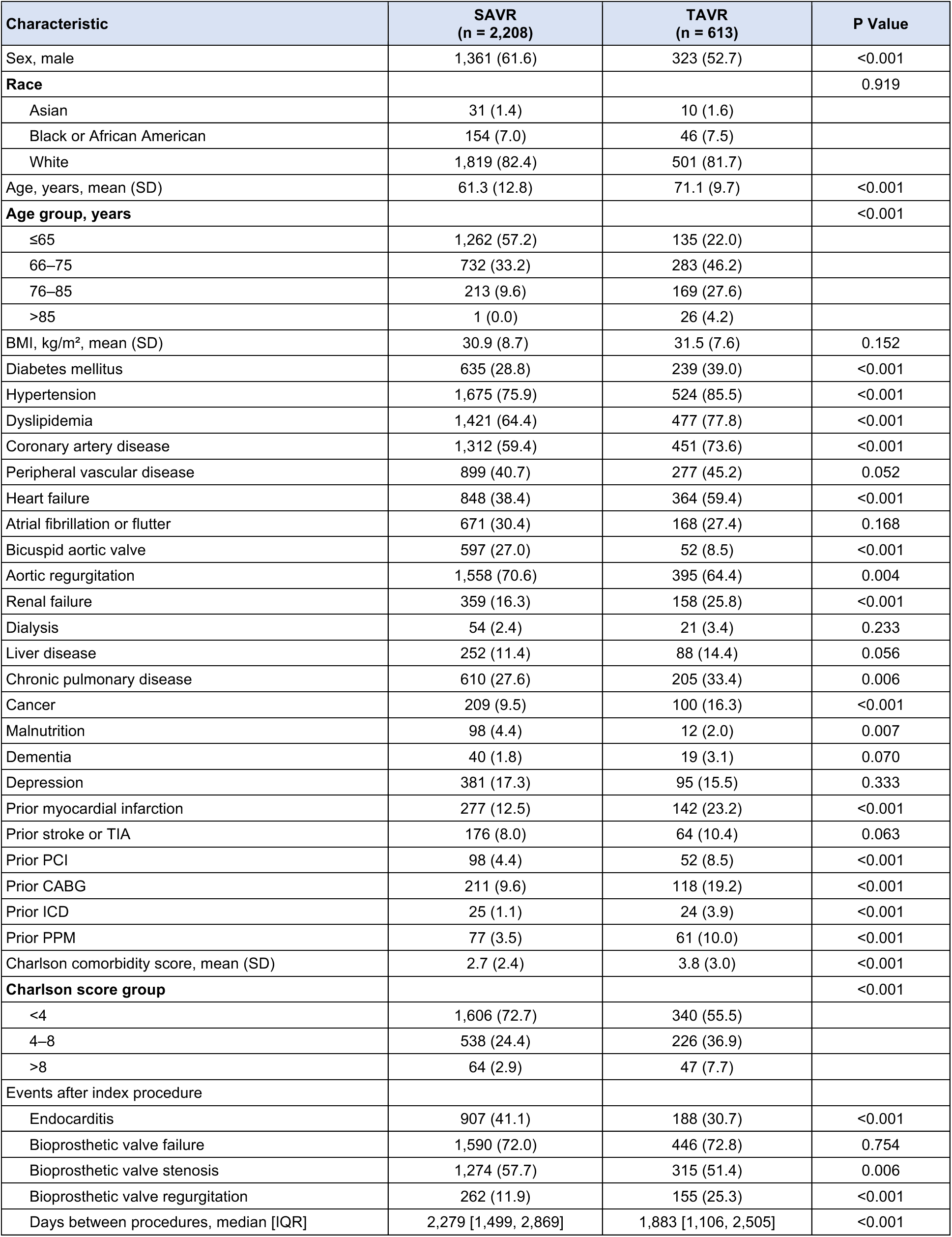
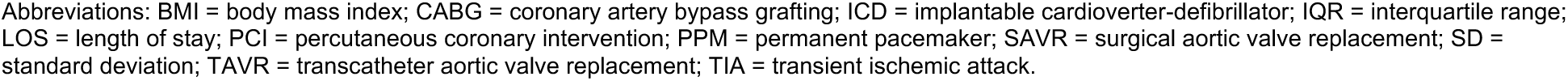
Characteristics of the Prespecified 1-Year Landmark Cohort Undergoing Repeat Aortic Valve Intervention.

| Characteristic | SAVR<br>(n = 2,208) | TAVR<br>(n = 613) | P Value |
| --- | --- | --- | --- |
| Sex, male | 1,361 (61.6) | 323 (52.7) | <0.001 |
| **Race** |  |  | 0.919 |
| Asian | 31 (1.4) | 10 (1.6) |  |
| Black or African American | 154 (7.0) | 46 (7.5) |  |
| White | 1,819 (82.4) | 501 (81.7) |  |
| Age, years, mean (SD) | 61.3 (12.8) | 71.1 (9.7) | <0.001 |
| **Age group, years** |  |  | <0.001 |
| ≤65 | 1,262 (57.2) | 135 (22.0) |  |
| 66–75 | 732 (33.2) | 283 (46.2) |  |
| 76–85 | 213 (9.6) | 169 (27.6) |  |
| >85 | 1 (0.0) | 26 (4.2) |  |
| BMI, kg/m<sup>2</sup>, mean (SD) | 30.9 (8.7) | 31.5 (7.6) | 0.152 |
| Diabetes mellitus | 635 (28.8) | 239 (39.0) | <0.001 |
| Hypertension | 1,675 (75.9) | 524 (85.5) | <0.001 |
| Dyslipidemia | 1,421 (64.4) | 477 (77.8) | <0.001 |
| Coronary artery disease | 1,312 (59.4) | 451 (73.6) | <0.001 |
| Peripheral vascular disease | 899 (40.7) | 277 (45.2) | 0.052 |
| Heart failure | 848 (38.4) | 364 (59.4) | <0.001 |
| Atrial fibrillation or flutter | 671 (30.4) | 168 (27.4) | 0.168 |
| Bicuspid aortic valve | 597 (27.0) | 52 (8.5) | <0.001 |
| Aortic regurgitation | 1,558 (70.6) | 395 (64.4) | 0.004 |
| Renal failure | 359 (16.3) | 158 (25.8) | <0.001 |
| Dialysis | 54 (2.4) | 21 (3.4) | 0.233 |
| Liver disease | 252 (11.4) | 88 (14.4) | 0.056 |
| Chronic pulmonary disease | 610 (27.6) | 205 (33.4) | 0.006 |
| Cancer | 209 (9.5) | 100 (16.3) | <0.001 |
| Malnutrition | 98 (4.4) | 12 (2.0) | 0.007 |
| Dementia | 40 (1.8) | 19 (3.1) | 0.070 |
| Depression | 381 (17.3) | 95 (15.5) | 0.333 |
| Prior myocardial infarction | 277 (12.5) | 142 (23.2) | <0.001 |
| Prior stroke or TIA | 176 (8.0) | 64 (10.4) | 0.063 |
| Prior PCI | 98 (4.4) | 52 (8.5) | <0.001 |
| Prior CABG | 211 (9.6) | 118 (19.2) | <0.001 |
| Prior ICD | 25 (1.1) | 24 (3.9) | <0.001 |
| Prior PPM | 77 (3.5) | 61 (10.0) | <0.001 |
| Charlson comorbidity score, mean (SD) | 2.7 (2.4) | 3.8 (3.0) | <0.001 |
| **Charlson score group** |  |  | <0.001 |
| <4 | 1,606 (72.7) | 340 (55.5) |  |
| 4–8 | 538 (24.4) | 226 (36.9) |  |
| >8 | 64 (2.9) | 47 (7.7) |  |
| **Events after index procedure** |  |  |  |
| Endocarditis | 907 (41.1) | 188 (30.7) | <0.001 |
| Bioprosthetic valve failure | 1,590 (72.0) | 446 (72.8) | 0.754 |
| Bioprosthetic valve stenosis | 1,274 (57.7) | 315 (51.4) | 0.006 |
| Bioprosthetic valve regurgitation | 262 (11.9) | 155 (25.3) | <0.001 |
| Days between procedures, median [IQR] | 2,279 [1,499, 2,869] | 1,883 [1,106, 2,505] | <0.001 |

Abbreviations: BMI = body mass index; CABG = coronary artery bypass grafting; ICD = implantable cardioverter-defibrillator; IQR = interquartile range; LOS = length of stay; PCI = percutaneous coronary intervention; PPM = permanent pacemaker; SAVR = surgical aortic valve replacement; SD = standard deviation; TAVR = transcatheter aortic valve replacement; TIA = transient ischemic attack.

The indications for repeat intervention differed between treatment groups. Endocarditis during follow-up was more frequently observed among patients with prior SAVR than among those with prior TAVR (41.1% vs. 30.7%; P<0.001), whereas the prevalence of bioprosthetic valve failure was similar between the two groups (72.0% vs. 72.8%; P=0.754). Kaplan-Meier analysis similarly demonstrated earlier repeat intervention following TAVR (Figure 5). Restricted mean survival time favored surgical bioprosthetic valves, providing an additional 4.5 months free from repeat intervention at 5 years (95% CI 3.1–5.9 months; P<0.001) and 10.7 months at 10 years (95% CI 8.1–13.3 months; P<0.001). To determine whether the observed difference was driven by infective endocarditis, we performed a prespecified subgroup analysis limited to patients with bioprosthetic valve failure in the absence of endocarditis. The results remained consistent. Compared with TAVR, surgical bioprosthetic valves provided an additional 5.4 months free from clinically significant valve failure requiring repeat intervention at 5 years (95% CI, 3.7–7.1 months; P<0.001) and 12.0 months at 10 years (95% CI, 8.5–15.5 months; P<0.001) (Supplemental Figure 3). Following propensity-score matching (Supplemental Table 4), the findings remained consistent. RMST continued to favor SAVR, with 2.3 additional months free from repeat intervention at 5 years (95% CI 0.3–4.2 months; P=0.024) and 4.0 additional months at 10 years (95% CI 0.4–7.7 months; P=0.031) (Supplemental Figure 4).

**Figure 5.**
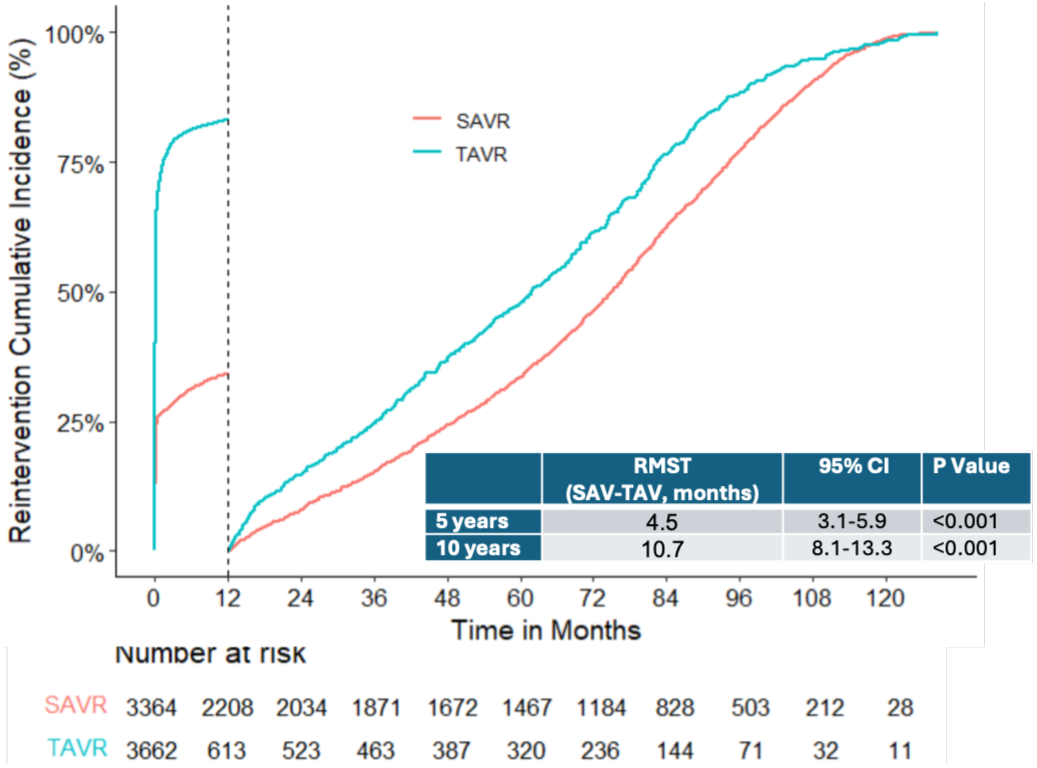
Prespecified 1-year landmark analysis of clinically significant bioprosthetic valve failure requiring repeat intervention following transcatheter (TAVR) and surgical aortic valve replacement (SAVR). Restricted mean survival time analysis demonstrated longer freedom from clinically significant valve failure requiring repeat intervention after SAVR than after TAVR. SAV = Surgical aortic valve; TAV = Transcatheter aortic valve.

On multivariable analysis, older age, male sex, higher body mass index, heart failure, chronic kidney disease, malignancy, malnutrition, dementia, depression, previous myocardial infarction, and prior stroke or transient ischemic attack were independently associated with a lower likelihood of repeat TAVR intervention beyond one year (Table 4).

**Table 4.** Multivariable Predictors of Clinically Significant Valve Failure Requiring Repeat Intervention Beyond 1 Year After Index TAVR.

|  |  | 95% CI |  |  |
| --- | --- | --- | --- | --- |
|  | OR | Lower | Upper | P Value |
| Sex, Male | 0.70 | 0.60 | 0.83 | <0.001 |
| BMI | 0.98 | 0.97 | 1.00 | 0.011 |
| Age | 0.92 | 0.92 | 0.93 | <0.001 |
| CHF | 0.82 | 0.69 | 0.98 | 0.028 |
| Renal Failure | 0.57 | 0.47 | 0.70 | <0.001 |
| Cancer | 0.77 | 0.61 | 0.95 | 0.017 |
| Malnutrition | 0.33 | 0.17 | 0.56 | <0.001 |
| Dementia | 0.44 | 0.27 | 0.68 | <0.001 |
| Depression | 0.70 | 0.56 | 0.88 | 0.003 |
| Prior Myocardial infarction | 0.75 | 0.61 | 0.93 | 0.009 |
| Prior Stroke/TIA | 0.75 | 0.57 | 0.97 | 0.036 |
| Prior CABG | 1.87 | 1.49 | 2.33 | 0.004 |
| Postprocedural acute kidney injury | 2.94 | 2.19 | 3.87 | <0.001 |
| Postprocedural major bleeding | 2.67 | 2.16 | 3.29 | <0.001 |

Abbreviations: BMI = body mass index; CABG = coronary artery bypass grafting; TAVR = transcatheter aortic valve replacement; TIA = transient ischemic attack.

## Discussion

In this contemporary national study of more than 300,000 patients undergoing TAVR, we provide a comprehensive evaluation of procedural outcomes, aortic valve reintervention, and clinically significant valve failure requiring repeat intervention. Five principal findings emerge. First, TAVR utilization increased dramatically during the study period and was accompanied by substantial improvements in early clinical outcomes. Second, repeat aortic valve intervention after TAVR remained uncommon and became progressively less frequent in contemporary practice. Third, redo-TAVR and surgical explantation represented distinct clinical pathways, with redo-TAVR performed predominantly during the early postprocedural period and surgical explantation occurring later. Fourth, redo-TAVR was associated with substantially lower perioperative morbidity and mortality, whereas surgical explantation was associated with superior long-term survival among propensity-matched hospital survivors. Finally, among patients undergoing repeat intervention beyond one year, clinically significant valve failure requiring repeat intervention occurred earlier after TAVR than after surgical bioprosthetic valve replacement.

Our findings demonstrate the remarkable evolution of TAVR over the past decade. As procedural volume increased more than tenfold, early clinical outcomes improved substantially, with significant reductions in mortality, bleeding, acute kidney injury, permanent pacemaker implantation, and overall MACE. These improvements likely reflect multiple advances occurring simultaneously, including refinements in transcatheter valve design, improved patient selection, greater operator experience, advances in preprocedural imaging, and increasing procedural standardization. Similar temporal improvements have been reported in national registries and randomized trials, supporting the continued maturation of TAVR as the dominant treatment strategy for severe aortic stenosis across a broad range of surgical risk (5,12–14).

As TAVR expands into younger populations, concerns regarding long-term valve performance have become increasingly important. In our study, however, repeat aortic valve intervention remained uncommon, with a cumulative incidence of only 2.7% at 10 years, and became progressively less frequent across successive procedural eras. These findings are consistent with contemporary national registry studies (15,16) and likely reflect improvements in valve technology, implantation techniques, and procedural planning. The declining incidence of reintervention is reassuring and suggests that clinically significant valve failure requiring another invasive procedure remains uncommon in current practice.

An important observation is the markedly different timing of the two reintervention strategies. Nearly 80% of redo-TAVR procedures occurred within 30 days of the index procedure, whereas surgical explantation was typically performed months to years later. This temporal separation strongly suggests that these procedures frequently address different clinical scenarios (17). Early redo-TAVR is more likely to reflect procedural complications, including valve malposition, embolization, severe paravalvular regurgitation, or immediate prosthetic dysfunction, whereas later surgical explantation more often reflects prosthetic valve endocarditis, structural valve degeneration, or complex anatomical situations that preclude repeat transcatheter intervention.

The present study highlights the complementary roles of redo-TAVR and surgical explantation in the management of failed transcatheter valves. Despite being performed in older patients, redo-TAVR was associated with substantially lower rates of mortality, stroke, acute kidney injury, major bleeding, and shorter hospitalization. These findings are consistent with previous multicenter experiences demonstrating the lower procedural risk of repeat transcatheter intervention (8,15,18). However, this early advantage did not translate into superior long-term survival. After propensity-score matching, hospital survivors undergoing surgical explantation experienced significantly better long-term survival. Several mechanisms may contribute to this observation. Surgical explantation completely removes the failed transcatheter prosthesis and allows definitive correction of associated pathology, whereas redo-TAVR necessarily leaves the original prosthesis in situ and may increase the risk of patient–prosthesis mismatch, coronary obstruction, impaired coronary access, and future management challenges related to multiple transcatheter valve layers. At the same time, residual selection bias cannot be excluded because patients referred for surgical explantation are generally younger, less frail, and considered suitable operative candidates despite statistical adjustment. Rather than suggesting that one strategy is universally superior, our findings support individualized treatment selection by a multidisciplinary heart team, incorporating patient age, comorbidity burden, anatomical considerations, mechanism of valve failure, and anticipated future treatment options.

One of the most clinically relevant findings of the present study is that, among patients surviving beyond one year and ultimately requiring repeat intervention, clinically significant valve failure requiring repeat intervention occurred earlier following TAVR than after surgical bioprosthetic valve replacement. Importantly, this finding should not be interpreted as direct evidence that transcatheter bioprosthetic valves have inferior intrinsic structural durability(19). Epic Cosmos does not capture serial echocardiographic measurements, valve hemodynamics, or imaging evidence of structural valve deterioration, precluding direct assessment of intrinsic valve performance. Instead, our analysis evaluates a patient-centered clinical endpoint—the occurrence of valve dysfunction severe enough to require another invasive procedure. Repeat intervention reflects the combined influence of structural valve degeneration, prosthetic valve endocarditis, valve thrombosis, paravalvular regurgitation, anatomical suitability for reintervention, competing mortality, physician decision-making, and patient preference. Accordingly, it represents an important measure of long-term clinical performance but should not be considered synonymous with intrinsic structural valve durability. Our findings therefore complement randomized clinical trials and registry studies by providing contemporary real-world estimates of freedom from clinically significant valve failure requiring repeat intervention in routine clinical practice (20–23).

The apparent discrepancy between the lower overall cumulative incidence of repeat intervention after TAVR and the earlier occurrence of clinically significant valve failure in the landmark analysis warrants consideration. Patients undergoing TAVR were substantially older and experienced higher competing mortality than those undergoing SAVR, reducing the opportunity to observe late repeat intervention. Furthermore, many redo-TAVR procedures occurred within the first postoperative year and likely reflected procedural complications rather than progressive valve degeneration. By restricting the analysis to patients surviving beyond one year, the landmark analysis was designed to better isolate late clinically significant valve failure while minimizing the influence of early technical failures and competing mortality. Together, these complementary analyses provide a more nuanced assessment of long-term valve performance than either approach alone.

### Study Limitations

This study has several limitations. First, its retrospective observational design precludes causal inference despite adjustment for measured confounders and propensity-score matching. Second, identification of procedures and clinical events relied on administrative coding, introducing the possibility of misclassification. Third, Epic Cosmos lacks detailed procedural and anatomical information, including valve type, implant depth, annular dimensions, coronary anatomy, and serial echocardiographic or computed tomography measurements, all of which influence procedural outcomes and long-term valve performance. Most importantly, clinically significant valve failure was defined using repeat aortic valve intervention rather than direct imaging evidence of structural valve deterioration. Repeat intervention is influenced not only by valve performance but also by patient life expectancy, competing mortality, anatomical suitability, physician judgment, and patient preference. In addition, reintervention strategies differ according to the index valve type: failed SAVR may be treated with transcatheter valve-in-valve implantation, whereas failed TAVR may require TAVR-in-TAVR or surgical explantation. Differences in the feasibility, risk, and clinical threshold for these interventions may therefore influence the likelihood that valve dysfunction is captured as a repeat intervention and could affect comparisons of clinical durability between TAVR and SAVR. Fourth, same-day reinterventions were excluded by design and therefore may have led to underestimation of early technical failures and 30-day reintervention and major adverse cardiovascular event rates in both treatment groups. Finally, the TAVR cohort spanned a period of substantial procedural evolution, with markedly increasing TAVR volume over time. Consequently, estimates extending to 10 years are necessarily driven predominantly by patients treated with earlier-generation transcatheter valves and may not be fully generalizable to contemporary valve platforms or younger patients with longer life expectancy. Future prospective studies incorporating serial multimodality imaging and standardized definitions of structural valve deterioration are needed to define the biological durability of contemporary transcatheter and surgical bioprosthetic valves.

## Conclusion

Contemporary TAVR demonstrates excellent procedural safety and a very low incidence of clinically significant valve failure requiring repeat intervention. Although redo-TAVR offers lower perioperative risk, surgical explantation was associated with superior long-term survival among selected hospital survivors. Furthermore, among patients surviving beyond one year after the index procedure, surgical bioprosthetic valves demonstrated greater clinical durability, reflected by longer freedom from clinically significant bioprosthetic valve failure requiring repeat intervention, than transcatheter valves. These findings provide contemporary real-world evidence to guide lifetime management of patients undergoing transcatheter aortic valve replacement.

## Abbreviations

CI: confidence interval
HR: hazard ratio
MACE: major adverse cardiovascular events
OR: odds ratio
RMST: restricted mean survival time
SAVR: surgical aortic valve replacement
TAVR: transcatheter aortic valve replacement

## Data Availability

All data produced are available online at https://cosmos.epic.com

## Central Illustration

Long-Term Reintervention, Clinical Valve Failure, and Outcomes After Transcatheter Aortic Valve Replacement. SAVR = surgical aortic valve replacement; TAVR = transcatheter aortic valve replacement.

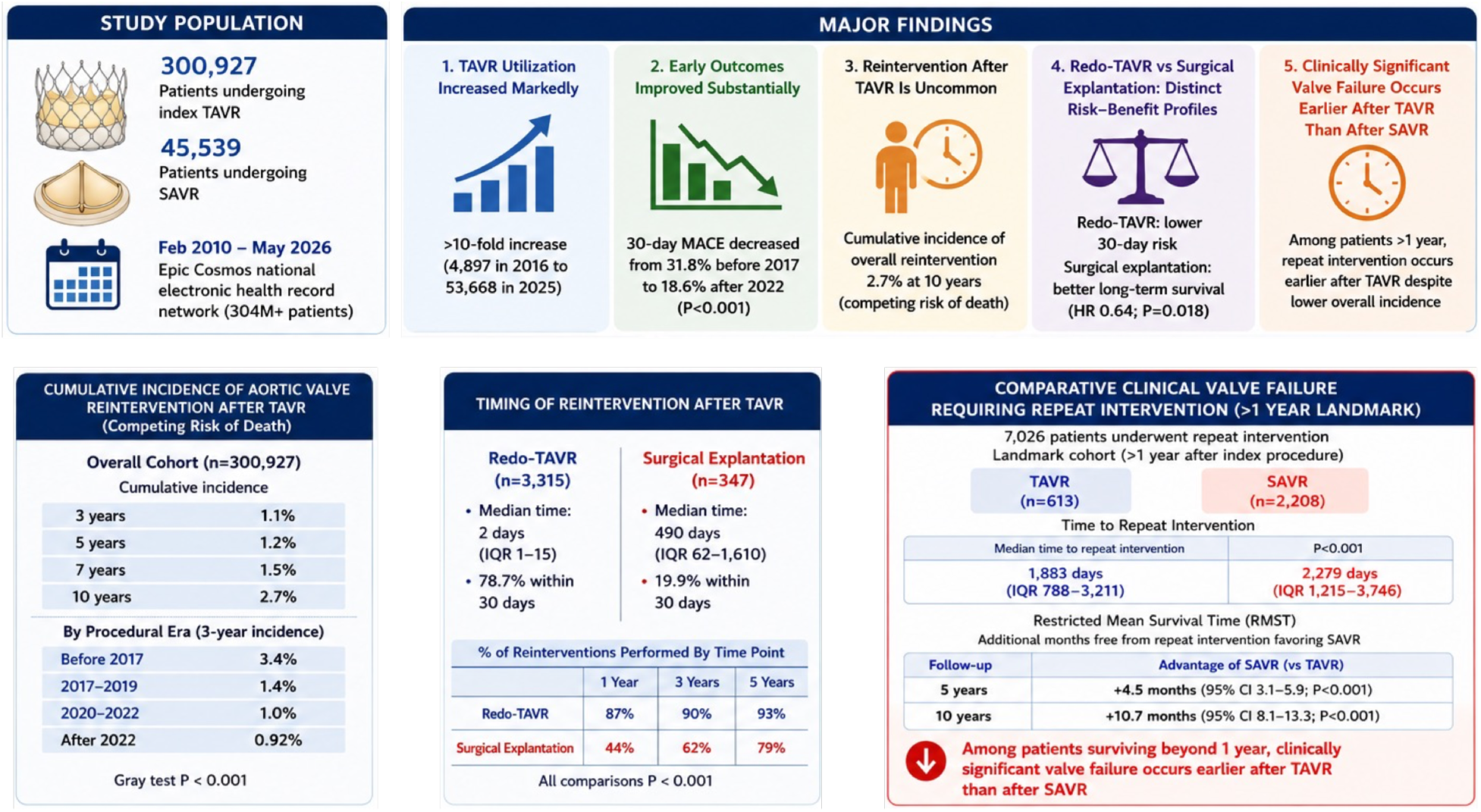

## Notes

### Competing Interest Statement

The authors have declared no competing interest.

### Author Declarations

Ethics committee/IRB of St Luke's University Health Network waived ethical approval for this work (IRB-FY2026-117).

