## Supplemental Data for "Long-Term Reintervention, Clinical Valve Failure, and Outcomes After Transcatheter Aortic Valve Replacement: A National Real-World Study"

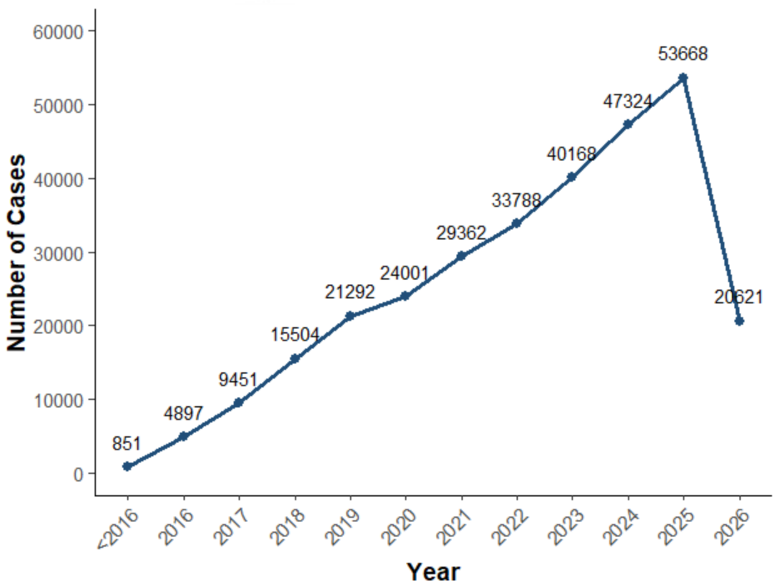


Supplemental Figure 1. Annual transcatheter aortic valve replacement (TAVR) procedural volume. Data for 2026 are presented through May 2026 only.

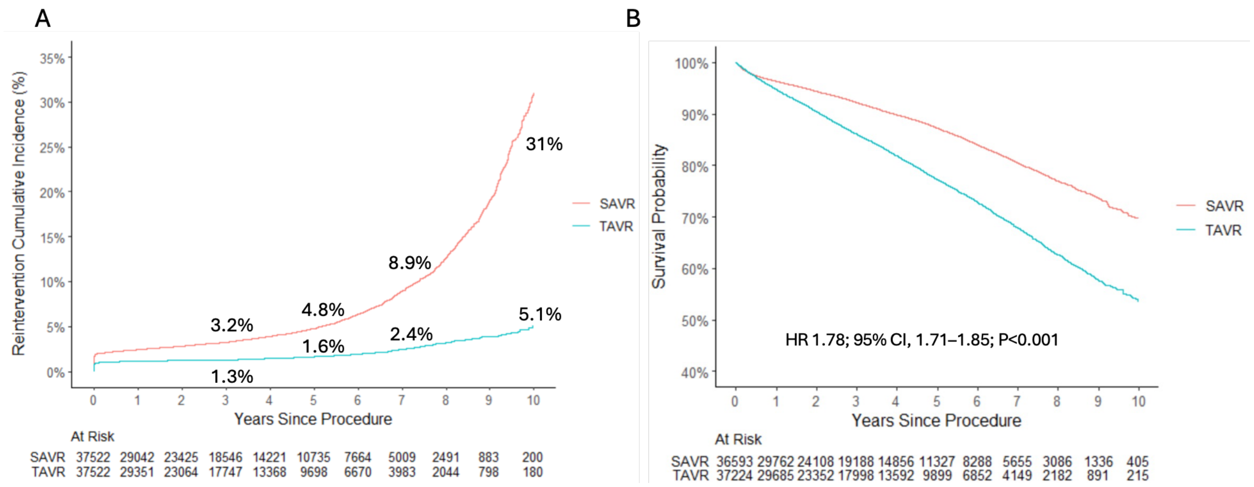


Supplemental Figure 2. Cumulative incidence of repeat aortic valve intervention and long-term survival after propensity-score matching of patients undergoing index transcatheter (TAVR) or surgical aortic valve replacement (SAVR). (A) Cumulative incidence of repeat aortic valve intervention after index TAVR and SAVR. (B) Kaplan–Meier survival curves among propensity-score–matched hospital survivors.

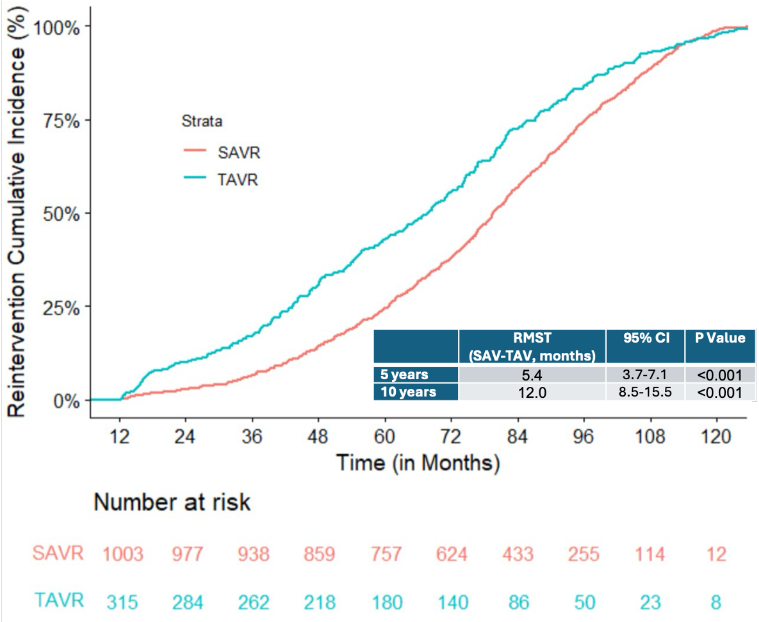


Supplemental Figure 3. Prespecified 1-year landmark analysis of clinical durability, defined as freedom from clinically significant bioprosthetic valve failure requiring repeat intervention, following transcatheter (TAVR) and surgical aortic valve replacement (SAVR) in patients with bioprosthetic valve failure in the absence of endocarditis. Restricted mean survival time analysis demonstrated longer freedom from clinically significant bioprosthetic valve failure requiring repeat intervention after SAVR than after TAVR. SAV = Surgical aortic valve; TAV = Transcatheter aortic valve.

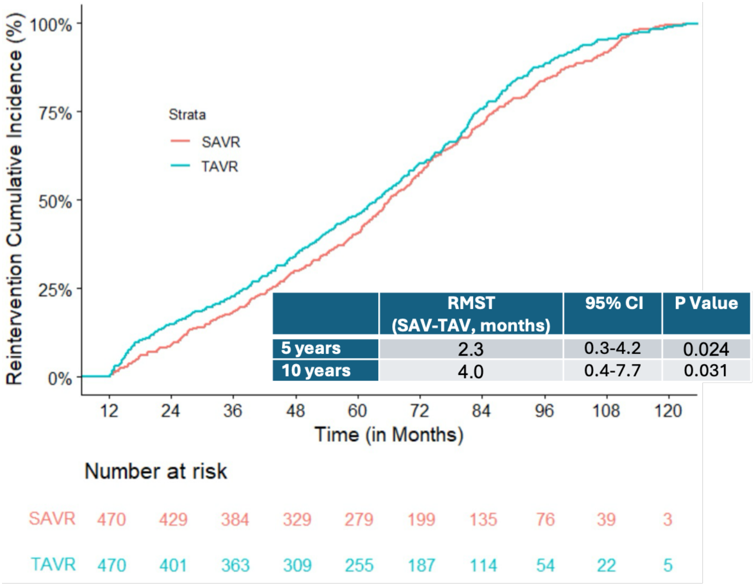


Supplemental Figure 4. Prespecified 1-year landmark analysis of clinical durability, defined as freedom from clinically significant bioprosthetic valve failure requiring repeat intervention, following transcatheter (TAVR) and surgical aortic valve replacement (SAVR) after propensity-score matching. Restricted mean survival time analysis demonstrated longer freedom from clinically significant bioprosthetic valve failure requiring repeat intervention after SAVR than after TAVR. SAV = Surgical aortic valve; TAV = Transcatheter aortic valve.

### Supplemental Table 1. ICD-10 diagnosis (CM), procedure (PCS) and CPT codes

| Characteristic | ICD-10 CM codes |
|---|---|
| **Comorbidities** | |
| Diabetes mellitus | E10.0, E10.1, E10.9, E11.0, E11.1, E11.9, E12.0, E12.1, E12.9, E13.0, E13.1, E13.9, E14.0, E14.1, E14.9, E10.2-E10.8, E11.2-E11.8, E12.2-E12.8, E13.2-E13.8, E14.2-E14.8 |
| Hypertension | I10.x, I11.x-I13.x, I15.x |
| Dyslipidemia | E78.x |
| Nicotine/tobacco use | F17.x, Z72.0, Z87.891 |
| Alcohol abuse | F10, E52, G62.1, I42.6, K29.2, K70.0, K70.3, K70.9, T51.x, Z50.2, Z71.4, Z72.1 |
| Drug abuse | F11.x-F16.x, F18.x, F19.x, Z71.5. Z72.2 |
| Obesity | E66.x |
| Coronary artery disease | I25.x |
| Atrial Fibrillation/Flutter | I48.x |
| Bicuspid aortic valve | Q23.0x, Q23.1x, Q23.8x, Q23.9x |
| Endocarditis | I33.x, I38, I39 |
| Bioprosthetic valve failure | T82.01XA, T82.02XA, T82.03XA, T82.09XA, T82.221A, T82.222A, T82.223A, T82.228A, T82.857A, T82.867A |
| Bioprosthetic valve stenosis | T82.857A |
| Bioprosthetic valve regurgitation | T82.03XA, T82.223A |
| Peripheral vascular disease | I70.x, I71.x, I73.1, I73.8, I73.9, I77.1, I79.0, I79.2, K55.1, K55.8, K55.9, Z95.8, Z95.9 |
| Congestive heart failure | I09.9, I11.0, I13.0, I13.2, I25.5, I42.0, 142.5-I42.9, I43.x, I50.x, P29.0 |
| Renal failure | I12.0, I13.1, N18.x, N19.x, N25.0, Z49.0-Z49.2, Z94.0, Z199.2 |
| Dialysis dependent | Z99.2 |
| Liver disease | B18.x, I85.x, I86.4, I98.2, K70.x, K71.1, K71.3-K71.5, K71.7, K72.x-K74.x, K76.0, K76.2-K76.9. Z94.4 |
| Chronic pulmonary disease | I27.8, 127.9, J40.x-J47.x, J60.x-J67.x, J68.4, J70.1, J70.3 |
| Obstructive sleep apnea | G47.33 |
| Coagulopathy | D65-D68.x, D69.1, D69.3-D69.6 |
| Cancer | C0x.x, C1x.x, C2x.x, C30.x, C31.x, C32.x, C33.x, C34.x, C37.x, C38.x, C39.x, C40.x, C41.x, C43.x, C45.x, C46.x, C47.x, C48.x, C49.x, C50, C51-58.x, C60-63.x, C76.x, C80.1, C81.x, C82.x, C83.x, C84.x, C85.x, C88.x, C9x.x |
| Malnutrition | E43, E44.x, E45, E46 |
| Dementia | F01.x, F02.x, F03.x, F04, F05, F06.1, F06.8, G13.2, G13.8, G30.x, G31.0x, G31.1, G31.2, G91.4, G94, R41.81, R54 |
| Depression | F20.4, F31.3-F31.5, F32.x, F33.x, F34.1, F41.2, F43.2 |
| **Previous history** | |
| Myocardial infarction | I25.2 |
| Stroke/TIA | Z86.73 |
| Cardiac arrest | Z86.74 |
| PCI | Z98.61, Z95.5 |
| CABG | Z95.1 |
| ICD | Z95.810 |
| PPM | Z95.0 |
| **In-hospital outcomes** | |
| Stroke | I63.x, I67.81, I67.82, G45.x, G46.x, H34.0x, H34.1x, H34.2x, I60.x, I61.x, I62.x, I97.820, I97.810 |
| Acute kidney injury | N17.x, N99.0 |
| Major bleeding | I97.610, I97.410, I97.618, I97.418, L76.22, L76.02, I97.51, L76.12, K92.0, K92.1, K92.2, K91.841, K91.62, R31.0, N99.821, N99.62, R04.x, J95.831, J95.62, R58, D62 |
| Heart block | I44.x |
| Complete heart block | I44.2 |

| Characteristic | ICD-10 CM codes |
| --- | --- |
| **Procedures** | **ICD-10 PCS /CPT codes** |
| TAVR | 02RF37H, 02RF38H, 02RF3JH, 02RF3KH, X2RF332, 02RF37Z, 02RF38Z, 02RF3JZ, 02RF3KZ/<br>33361, 33362, 33363, 3364, 33365, 33366, 33367, 33368, 33369, 33378 |
| Surgical AVR | 02RF07Z, 02RF08Z, 02RF0JZ, 02RF0KZ/<br>33405, 33406 |
| PPM | 0JH604Z, 0JH605Z, 0JH606Z, 0JH607Z/<br>33206, 33207, 33208 |

### Supplemental Table 2. Baseline Characteristics and Clinical Outcomes of Patients Undergoing Repeat Intervention After Index TAVR

| Characteristic | TAVR→TAVR (n = 3,315) | TAVR→SAVR (n = 347) | P Value |
|---|---|---|---|
| Sex, male | 1,865 (56.3) | 196 (56.5) | 0.981 |
| **Race** | | | 0.728 |
| Asian | 52 (1.6) | 9 (2.6) | |
| Black or African American | 210 (6.3) | 22 (6.3) | |
| White | 2,705 (81.6) | 282 (81.3) | |
| **Age group, years** | | | <0.001 |
| ≤65 | 280 (8.4) | 74 (21.3) | |
| 66–75 | 801 (24.2) | 160 (46.1) | |
| 76–85 | 1,427 (43.0) | 106 (30.5) | |
| >85 | 807 (24.3) | 7 (2.0) | |
| Age, years, mean (SD) | 78.8 (9.1) | 70.9 (9.2) | <0.001 |
| Age at Index TAVR, mean (SD) | 78.1 (9.2) | 68.4 (9.7) | <0.001 |
| BMI, kg/m², mean (SD) | 30.2 (6.5) | 30.5 (7.5) | 0.935 |
| Diabetes mellitus | 1,552 (46.8) | 179 (51.6) | 0.102 |
| Hypertension | 3,195 (96.4) | 332 (95.7) | 0.609 |
| Dyslipidemia | 2,998 (90.4) | 308 (88.8) | 0.364 |
| Coronary artery disease | 2,975 (89.7) | 306 (88.2) | 0.416 |
| Peripheral vascular disease | 2,017 (60.8) | 241 (69.5) | 0.002 |
| Heart failure | 2,842 (85.7) | 302 (87.0) | 0.562 |
| Atrial fibrillation or flutter | 1,620 (48.9) | 206 (59.4) | <0.001 |
| Bicuspid aortic valve | 243 (7.3) | 71 (20.5) | <0.001 |
| Renal failure | 1,776 (53.6) | 164 (47.3) | 0.029 |
| Dialysis | 194 (5.9) | 22 (6.3) | 0.805 |
| Liver disease | 634 (19.1) | 93 (26.8) | 0.001 |
| Chronic pulmonary disease | 1,502 (45.3) | 130 (37.5) | 0.006 |
| Cancer | 831 (25.1) | 79 (22.8) | 0.380 |
| Malnutrition | 375 (11.3) | 50 (14.4) | 0.104 |
| Dementia | 578 (17.4) | 49 (14.1) | 0.138 |
| Prior myocardial infarction | 1,379 (41.6) | 126 (36.3) | 0.065 |
| Prior stroke or TIA | 757 (22.8) | 83 (23.9) | 0.697 |
| Prior PCI | 490 (14.8) | 33 (9.5) | 0.010 |
| Prior CABG | 698 (21.1) | 91 (26.2) | 0.031 |
| Prior ICD | 213 (6.4) | 25 (7.2) | 0.656 |
| Prior PPM | 714 (21.5) | 73 (21.0) | 0.883 |
| **Events after index procedure** | | | |
| Endocarditis | 141 (4.3) | 119 (34.3) | <0.001 |
| Bioprosthetic valve failure | 400 (12.1) | 171 (49.3) | <0.001 |
| Bioprosthetic valve stenosis | 255 (7.7) | 114 (32.9) | <0.001 |
| Bioprosthetic valve regurgitation | 154 (4.6) | 59 (17.0) | <0.001 |
| **In-hospital procedural outcomes** | | | |
| Stroke | 43 (1.3) | 21 (6.1) | <0.001 |
| Acute kidney injury | 116 (3.5) | 72 (20.7) | <0.001 |
| Major bleeding | 158 (4.8) | 143 (41.2) | <0.001 |

| Characteristic | TAVR→TAVR (n = 3,315) | TAVR→SAVR (n = 347) | P Value |
|---|---|---|---|
| Heart block | 378 (11.4) | 70 (20.2) | <0.001 |
| Third-degree heart block | 123 (3.7) | 41 (11.8) | <0.001 |
| PPM implantation | 240 (7.2) | 40 (11.5) | 0.006 |
| Days between procedures, median [IQR] | 2.0 [1.0, 15.0] | 490.0 [62.0, 1,610.0] | <0.001 |
| Length of hospital stay, days, median [IQR] | 3.0 [1.0, 8.0] | 12.0 [7.0, 20.0] | <0.001 |
| Major adverse cardiovascular events | 557 (16.8) | 207 (59.7) | <0.001 |
| 30-day mortality | 107 (3.2) | 31 (8.9) | <0.001 |
| 1-year mortality | 349 (10.5) | 49 (14.1) | 0.051 |
| Reintervention within 30 days | 2,609 (78.7) | 69 (19.9) | <0.001 |
| Reintervention within 90 days | 2,783 (84.0) | 105 (30.3) | <0.001 |

Abbreviations: BMI = body mass index; CABG = coronary artery bypass grafting; ICD = implantable cardioverter-defibrillator; IQR = interquartile range; LOS = length of stay; PCI = percutaneous coronary intervention; PPM = permanent pacemaker; SAVR = surgical aortic valve replacement; SD = standard deviation; TAVR = transcatheter aortic valve replacement; TIA = transient ischemic attack.

### Supplemental Table 3. Baseline Characteristics of the Propensity Score-Matched Hospital Survivor Cohort Undergoing Repeat Intervention

| Characteristic | TAVR→TAVR (n = 273) | TAVR→SAVR (n = 273) | P Value |
|---|---|---|---|
| Sex, male | 146 (53.5) | 154 (56.4) | 0.547 |
| **Race** | | | — |
| Asian | 6 (2.2) | 7 (2.6) | |
| Black or African American | 15 (5.5) | 16 (5.9) | |
| White | 228 (83.5) | 221 (81.0) | |
| **Year of Procedure** | | | 0.997 |
| 2013 | 2 (0.7) | 1 (0.4) | |
| 2014 | 2 (0.7) | 2 (0.7) | |
| 2015 | 7 (2.6) | 9 (3.3) | |
| 2016 | 31 (11.4) | 29 (10.6) | |
| 2017 | 22 (8.1) | 27 (9.9) | |
| 2018 | 29 (10.6) | 34 (12.5) | |
| 2019 | 43 (15.8) | 44 (16.1) | |
| 2020 | 26 (9.5) | 22 (8.1) | |
| 2021 | 29 (10.6) | 30 (11.0) | |
| 2022 | 23 (8.4) | 25 (9.2) | |
| 2023 | 23 (8.4) | 19 (7.0) | |
| 2024 | 21 (7.7) | 16 (5.9) | |
| 2025 | 11 (4.0) | 12 (4.4) | |
| 2026 | 4 (1.5) | 3 (1.1) | |
| **Age group, years** | | | 0.989 |
| ≤65 | 56 (20.5) | 53 (19.4) | |
| 66–75 | 120 (44.0) | 123 (45.1) | |
| 76–85 | 92 (33.7) | 92 (33.7) | |
| >85 | 5 (1.8) | 5 (1.8) | |
| Age, years, mean (SD) | 71.3 (10.1) | 71.9 (8.0) | 0.450 |
| BMI, kg/m², mean (SD) | 30.6 (7.5) | 30.5 (7.6) | 0.901 |
| Diabetes mellitus | 134 (49.1) | 134 (49.1) | 1.000 |
| Hypertension | 258 (94.5) | 262 (96.0) | 0.547 |
| Dyslipidemia | 234 (85.7) | 241 (88.3) | 0.445 |
| Coronary artery disease | 242 (88.6) | 238 (87.2) | 0.694 |
| Peripheral vascular disease | 193 (70.7) | 185 (67.8) | 0.516 |
| Heart failure | 230 (84.2) | 234 (85.7) | 0.719 |
| Atrial fibrillation or flutter | 142 (52.0) | 152 (55.7) | 0.440 |
| Bicuspid aortic valve | 47 (17.2) | 48 (17.6) | 1.000 |
| Renal failure | 116 (42.5) | 121 (44.3) | 0.730 |
| Dialysis | 10 (3.7) | 11 (4.0) | 1.000 |
| Liver disease | 76 (27.8) | 64 (23.4) | 0.281 |
| Chronic pulmonary disease | 128 (46.9) | 110 (40.3) | 0.142 |
| Cancer | 52 (19.0) | 59 (21.6) | 0.523 |
| Malnutrition | 41 (15.0) | 35 (12.8) | 0.536 |
| Dementia | 46 (16.8) | 35 (12.8) | 0.229 |
| Prior myocardial infarction | 96 (35.2) | 99 (36.3) | 0.858 |
| Prior stroke or TIA | 64 (23.4) | 63 (23.1) | 1.000 |
| Prior PCI | 33 (12.1) | 28 (10.3) | 0.587 |
| Prior CABG | 68 (24.9) | 69 (25.3) | 1.000 |
| Prior ICD | 19 (7.0) | 20 (7.3) | 1.000 |

| Characteristic | TAVR→TAVR (n = 273) | TAVR→SAVR (n = 273) | P Value |
|---|---|---|---|
| Prior PPM | 54 (19.8) | 55 (20.1) | 1.000 |

Abbreviations: BMI = body mass index; CABG = coronary artery bypass grafting; ICD = implantable cardioverter-defibrillator; IQR = interquartile range; LOS = length of stay; PCI = percutaneous coronary intervention; PPM = permanent pacemaker; SAVR = surgical aortic valve replacement; SD = standard deviation; TAVR = transcatheter aortic valve replacement; TIA = transient ischemic attack.

### Supplemental Table 4. Baseline Characteristics of the Propensity Score-Matched 1-Year Landmark Cohort

| Characteristic | SAVR (n = 470) | TAVR (n = 470) | P Value |
|---|---|---|---|
| Sex, male | 254 (54.0) | 257 (54.7) | 0.896 |
| **Race** | | | 0.977 |
| Asian | 6 (1.3) | 7 (1.5) | |
| Black or African American | 30 (6.4) | 34 (7.2) | |
| White | 394 (83.8) | 389 (82.8) | |
| **Year of Procedure** | | | 0.999 |
| 2013 | 3 (0.6) | 3 (0.6) | |
| 2014 | 2 (0.4) | 2 (0.4) | |
| 2015 | 40 (8.5) | 41 (8.7) | |
| 2016 | 103 (21.9) | 92 (19.6) | |
| 2017 | 80 (17.0) | 82 (17.4) | |
| 2018 | 69 (14.7) | 72 (15.3) | |
| 2019 | 76 (16.2) | 78 (16.6) | |
| 2020 | 36 (7.7) | 38 (8.1) | |
| 2021 | 29 (6.2) | 27 (5.7) | |
| 2022 | 14 (3.0) | 16 (3.4) | |
| 2023 | 13 (2.8) | 14 (3.0) | |
| 2024 | 4 (0.9) | 4 (0.9) | |
| 2025 | 1 (0.2) | 1 (0.2) | |
| Age, years, mean (SD) | 69.4 (8.3) | 69.3 (9.3) | 0.839 |
| **Age group, years** | | | 0.655 |
| ≤65 | 120 (25.5) | 125 (26.6) | |
| 66–75 | 230 (48.9) | 236 (50.2) | |
| 76–85 | 119 (25.3) | 109(23.2) | |
| >85 | 1 (0.2) | 0 (0.0) | |
| BMI, kg/m², mean (SD) | 31.1 (6.3) | 31.6 (7.7) | 0.296 |
| Diabetes mellitus | 175 (37.2) | 176 (37.4) | 1.000 |
| Hypertension | 405 (86.2) | 400 (85.1) | 0.710 |
| Dyslipidemia | 358 (76.2) | 357 (76.0) | 1.000 |
| Coronary artery disease | 336 (71.5) | 334 (71.1) | 0.943 |
| Peripheral vascular disease | 202 (43.0) | 196 (41.7) | 0.741 |
| Heart failure | 262 (55.7) | 259 (55.1) | 0.896 |
| Atrial fibrillation or flutter | 127 (27.0) | 129 (27.4) | 0.942 |
| Bicuspid aortic valve | 48 (10.2) | 49 (10.4) | 1.000 |
| Aortic regurgitation | 305(64.9) | 306 (65.1) | 1.000 |
| Renal failure | 102 (21.7) | 103 (21.9) | 1.000 |
| Dialysis | 14 (3.0) | 13 (2.8) | 1.000 |
| Liver disease | 60 (12.8) | 66 (14.0) | 0.632 |
| Chronic pulmonary disease | 159 (33.8) | 155 (33.0) | 0.836 |
| Cancer | 72 (15.3) | 73 (15.5) | 1.000 |
| Malnutrition | 13 (2.8) | 9 (1.9) | 0.517 |
| Dementia | 12 (2.6) | 13 (2.8) | 1.000 |
| Prior myocardial infarction | 103 (21.9) | 96 (20.4) | 0.632 |
| Prior stroke or TIA | 43 (9.1) | 39 (8.3) | 0.729 |
| Prior PCI | 40 (8.5) | 34 (7.2) | 0.545 |
| Prior CABG | 74 (15.7) | 78 (16.6) | 0.790 |

| Characteristic | SAVR (n = 470) | TAVR (n = 470) | P Value |
|---|---|---|---|
| Prior ICD | 16 (3.4) | 16 (3.4) | 1.000 |
| Prior PPM | 32 (6.8) | 39 (8.3) | 0.459 |
| Charlson comorbidity score, mean (SD) | 3.5 (2.6) | 3.5 (2.9) | 0.981 |
| **Charlson score group** | | | 0.358 |
| <4 | 270 (57.4) | 283 (60.2) | |
| 4–8 | 176 (37.4) | 157 (33.4) | |
| >8 | 24 (5.1) | 30 (6.4) | |

Abbreviations: BMI = body mass index; CABG = coronary artery bypass grafting; ICD = implantable cardioverter-defibrillator; IQR = interquartile range; LOS = length of stay; PCI = percutaneous coronary intervention; PPM = permanent pacemaker; SAVR = surgical aortic valve replacement; SD = standard deviation; TAVR = transcatheter aortic valve replacement; TIA = transient ischemic attack.
